# Reduced mortality and shortened ICU stay in SARS-COV-2 patients: a low PEEP strategy

**DOI:** 10.1101/2020.05.03.20089318

**Authors:** Samuele Ceruti, Marco Roncador, Olivier Gié, Giovanni Bona, Martina Iattoni, Maira Biggiogero, Pier Andrea Maida, COVID-19 Clinical Management Team, Christian Garzoni, Romano Mauri, COVID-19 Clinical Management Team, Carola Biondi, Sara Canovi, Mattia Cavagna, Bruno Di Mari, Zsofia Horvath, Rosambra Leo, Giorgia Lo Presti, Giorgia Luvini, Dario Meloni, Francesca Orlando, Sara Ravasi, Roberta Sonzini, Giuseppe Tinessa

## Abstract

**Background:** Intensive Care Unit (ICU) management of COVID-19 patients with severe hypoxemia is associated with high mortality. We implemented a ‘care map’, as a standardized multidisciplinary approach to improve patients monitoring using: uniform patient selection for ICU admission, a low-PEEP strategy and a pharmacologic strategic thromboembolism management.

**Methods:** A standardized protocol for managing COVID-19 patients and ICU admissions was implemented through accurate Early Warning Score (EWS) monitoring and thromboembolism prophylaxis at hospital admission. Dyspnea, mental confusion or SpO_2_ less than 85% were criteria for ICU admission. Ventilation approach employed low PEEP values (about 10 cmH_2_O in presence of lung compliance > 40 mL/cmH_2_O) and FiO_2_ as needed. In presence of lower lung compliance (< 40 mL/cmH_2_O) PEEP value was increased to about 14 cmH_2_O.

**Findings:** From March 16^th^ to April 12^nd^ 2020, 41 COVID-19 patients were admitted to our ICU from a total of 310 patients. 83% (34) of them needed mechanical ventilation. The ventilation approach chosen employed low PEEP value based on BMI (PEEP 11± 3.8 (10-12) cmH_2_O if BMI < 30 Kg/m^2^; PEEP 15± 3.26 (12-18) cmH_2_O if BMI >30 Kg/m^2^). To date, ten patients (24%) died, four (9.7%) received mechanical ventilation, two were transferred to another hospital and 25 (60.9%) were discharged from ICU after a median of nine days.

**Interpretation:** A multimodal approach for COVID-19 patients is mandatory. The knowledge of this multi-organ disease is growing rapidly, requiring improvements in the standard of care. Our approach implements an accurate pre-ICU monitoring and strict selection for ICU admission, and allows to reduce mechanical ventilation, ICU stay and mortality.

**Funding:** No funding has been required.

## BACKGROUND

Severe acute respiratory syndrome coronavirus 2 (SARS-CoV-2) is the cause of COVID-19, a pandemic that has affected more than 3,000,000 individuals and caused nearly 200,000 deaths since initial detection of the virus up to the end of April 2020^1^.

Epidemiologic data from China and Italy underline the severity of the syndrome, with a critical load for intensive care units (ICU) and a high mortality ^2,3^. In this phase of the pandemic, detailed reports describing management for COVID-19 patients admitted to the ICU are relevant ^4^ for a better clinical characterization and for guiding decision making in the severe hypoxemia affecting these subjects ^5^. In particular, newer pathophysiological understanding of the disease, as reported by Cronin ^6^, Nieman ^7^, Gattinoni ^8^ and Bendjelid ^9^, are keys for a better clinical evaluation and management.

On the basis of this newer concepts of COVID-19, we structured a ‘care map’ based on three relevant aspects: standardized criteria selection for ICU admission, a strategic antithrombotic therapy and low PEEP strategy.

The aim of our report is to describe the achievement obtained, in terms of survival, ICU length-of-stay and duration of mechanical ventilation (MV), by a multidisciplinary intervention which required a strong collaboration between ICU and other Departments, comparing them with current data ^2,3^. We further describe the demographic characteristics, coexisting conditions, critical care management and outcomes among patients admitted to Clinica Luganese Moncucco (CLM) ICU during the first four weeks of the outbreak in Canton Ticino area.

## METHODS

### Study population and data

A retrospective analysis was conducted on all consecutive patients with acute respiratory distress for COVID-19 pneumonia admitted to ICU from March 16^th^ to April 12^nd^, 2020. All patients’ relevant data like demographics, severity score (NEMS - nine equivalents of nursing manpower use score -, SAPS - simplified acute physiology score -), clinical information and laboratory/radiological results were obtained during patient’s hospitalization from electronic health records. Standard biological workup included complete blood count, CRP, ferritin, ASAT, ALAT, ionogram, creatinine, urea, D-dimer, Prothrombin Time (PT), activated Partial Thromboplastin Time (aPTT), fibrinogen, blood gas analysis, SvO_2_, pro-BNP, blood and urines cultures, urine research for Legionella antigen. A specific workup included a pulmonary ultrasound, a chest x-ray, transthoracic and transesophageal echocardiography (to establish the global cardiac function before any pronation). A thoracic CT-scan was considered available if it has been performed during the stay in the Internal Medicine Department during the last 24 hours before the admission in the ICU.

### Care Map description

Indication for ICU admission and oro-tracheal intubation (OTI) was routinely established by the intensive care specialist or senior anesthesiologist on duty, according to the ‘care map’ based on standardized criteria selection, low PEEP strategy and pharmacologic antithrombotic management.

#### Criteria selection for ICU admission

Requests of counseling for ICU admission came from the Department of Internal Medicine and from the Emergency Department (ED). With the aim of quickly identifying the worsening of clinical conditions ^11,12^, all consultations were recorded by reporting patient’s symptoms, SpO_2_, blood gas analysis values (if available) and clinical decision for admission or not to the ICU. Patients presenting partial respiratory failure combining peripheral saturation (SpO_2_) lower than 85% and dyspnea (or mental confusion), or patients with dyspnea (or mental confusion) alone, were eligible to be admitted in ICU. Exclusion criteria were the will of the patient not to be intubated, cardiocirculatory arrest following hypoxia, metastatic oncological disease, end-stage neurodegenerative disease, severe and irreversible chronic disease (heart failure NYHA IV, COPD GOLD D, liver cirrhosis Child-Pugh > 8, severe dementia) ^10^. With the aim to avoid a misleading interpretation in ICU mortality, we decided to perform an extra evaluation on patients excluded from ICU, to ensure about their survival status.

#### Ventilation settings – low PEEP strategy -

After endotracheal intubation, we initially provided low PEEP-value strategy based on BMI (PEEP 10 cmH_2_O if BMI < 30 Kg/m^2^, PEEP 12 cmH_2_O if BMI 30-50 Kg/m^2^, PEEP 15 cmH_2_O if BMI > 50 Kg/m^2^), subsequently adjusted in case of hypoxia (PaO_2_ < 60 mmHg / 8 kPa) according to *ARDSnet PEEP table* ^13,14^; in addition, we used protective ventilation strategy (TV 6-8 ml/Kg, P_plat_ < 30 cmH_2_O) with permissive hypercapnia (pH > 7.20) according to standard care ^15^ and immediate pronation. A deep sedation was maintained during first 36 hours using Midazolam (and eventually Ketamine), adding a paralyzing agent (Rocuronium) during first 24 hours and subsequently just in case of patient-ventilator asynchrony. Drugs dosages have adapted to pursue a Richmond Agitation and Sedation Scale (RASS) of −4 and a Train-Of-Four (TOF) around 1/4.

### Pharmacologic antithrombotic management

Given the high risk of Deep Venous Thrombosis (DVT) and Pulmonary Embolism (PE) ^15^, as patients were admitted to ICU we started with a ‘liberal’ prophylactic anticoagulation (Enoxaparine 60 mg bid SC if weight > 80 Kg, Enoxaparine 40 mg bid SC if weight < 80 Kg, Unfractioned Heparin in case of acute kidney injury - AKI) associated with ultrasound Color-Doppler lower limbs daily monitoring. In case of plasmatic D-dimer level greater than 1’500 ng/ml or documented thrombosis, anticoagulation treatment was switched to a therapeutic dose (Enoxaparine 1 mg/Kg bid SC – Unfractioned Heparin in case of AKI at 14 UI/Kg/day in perfusion, adapted according to anti-Xa values), according to our ‘care map’ concerning the antithrombotic management.

### Patients Clinical Evolution

Intensive supportive care was managed according to the evolution of the inflammatory parameters (CRP, CK, LDH and ferritin) and the stability of the P/F-ratio after each supination during the following days. In case of a favorable evolution, sedation was reduced to RASS −3/−2 switching on Propofol. By improvement of blood oxygen levels, we proceeded to reduce FiO_2_ up to 35-40% FiO_2_ values without reducing PEEP. Once all the clinical and biological inflammatory parameters were constantly reduced for almost three consecutive days, patients were gently weaned from PEEP by keeping a PaO_2_ > 60 mmHg (8 kPa). The choice of removing the endotracheal tube was made by the doctor in charge according to usual standard of care.

Deep vein thrombosis, PE, Ventilator-Associated-Pneumonia (VAP) and Acute Kidney Injury (AKI) have been the main complications arose in patients admitted to ICU: DVTs and PEs were defined as *suspected* with an increase in serum D-dimer values over 1,500 ng/ml, while they were considered as *confirmed* by ultrasound or CT-scan positive finding, defined according to current clinical standards. VAPs were identified according to usual standard care ^16^ with an increase in secretions, in their quality and quantity, requiring an increase in the FiO_2_ administration ^17^. Each case of AKI RIFLE F requesting CVVHDF has also been reported. All complications, administered drugs and adverse events occurring during the stay in intensive care, were registered and reported in the electronic medical record.

### Statistical analysis and comparison with current literature

Descriptive statistics were used to summarize the clinical data collected. No statistical sample size calculation was performed. We present continuous measurements as mean (min-max, SD) otherwise as median (IQR) if they are normally distributed. Categorical variables were reported as counts and percentages. Test statistics and survival analysis were performed with R v.3.6.1 and the Kaplan-Meier estimator from CRAN “surv” package. Data was subsequently compared with a similar in number and follow-up cohort published by Bhatraju et al^18^ - complete patient data was retrieved from the supplementary appendix.

### Ethics Committee permissions

This study has been notified to the Ethics Committees of Canton Ticino. According to the local Federal rules, it has been approved as a clinical data collection case series.

### Role of the funding source

No funding has been required. The corresponding author confirms that he had full access to all the data in the study and he had final responsibility for the decision to submit for publication.

## RESULTS

### Pre-ICU patients’ evaluation

310 patients with COVID-19 symptoms presented to our Clinic. According to exclusion criteria, 54 of them were not admitted to the ICU as they had a “do not resuscitate” order (DNR) in place before hospital admission (Figure 1); on April 12^th^, 14 of DNR patients (25%) died. Globally, 41 critically ill patients with laboratory-confirmed SARS-CoV-2 infection were admitted to the ICU (Figure 1); 10 patients (24%) has been admitted directly from the ED, 5 (12%) from others hospitals and 26 (64%) from internal consultation (Figure 1) Demographic and clinical characteristics of patients were shown in Table 1. Patients had a mean age of 64±16·5 years; most of them were men, often burdened by one or more chronic medical conditions (Table 1). At ICU admission, most patients showed hemodynamic stability. A chest CT-scan was obtained in 23 (56%) patients; all of them showed bilateral ground glass opacities and four of them showed consolidations in addition (Table 1).

**Figure 1.**
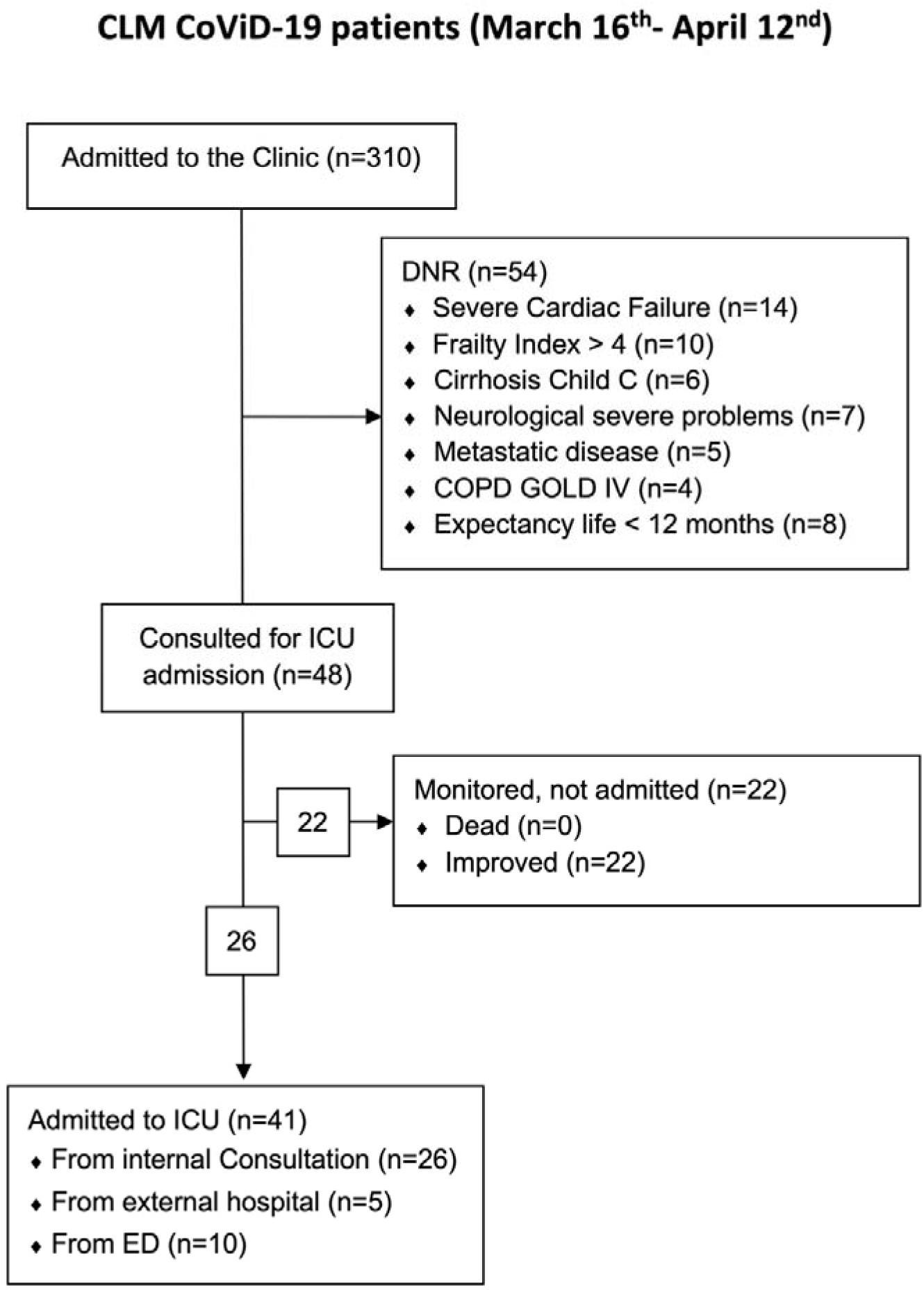
Management of the CoViD-19 patients evaluated at the CLM.

**Table 1.** Baseline characteristics.

|  | Unit | n.v. | Results |
| --- | --- | --- | --- |
| **DEMOGRAPHIC DATA:** |  |  |  |
| Patients admitted to ICU | n |  | 41 |
| Age | years |  | 64 ± 16.5 (29–85) |
| Male | n |  | 35 (85%) |
| BMI | kg/m<sup>2</sup> |  | 28.4 ± 4.94 (19.0–41.1) |
| SAPS |  |  | 45 ± 18.40 (13–94) |
| NEMS |  |  | 30 ± 10.4 (9–42) |
| **COMORBIDITIES:** |  |  |  |
| Arterial Hypertension | n |  | 19 (46%) |
| Ischemic cardiopathy | n |  | 6 (14.6%) |
| Diabetes | n |  | 15 (36.5%) |
| Obstructive Sleep Apnea Syndrome | n |  | 6 (14.6%) |
| COPD | n |  | 4 (9.7%) |
| **AT ADMISSION** |  |  |  |
| **HEMODYNAMICS:** |  |  |  |
| Mean duration of symptoms | days |  | 7 (1–30) |
| Systolic arterial pressure | mmHg | 110-140 | 129 (80–180) |
| Diastolic arterial pressure | mmHg | 60-80 | 67 (40–110) |
| Heart Rate | bpm | 60-100 | 88 (55–160) |
| Temperature | °C | 36 – 38.3 | 37.0 (35.6–39) |
| Lactate | mmol/L | < 2.0 | 1.5 (0.5–2.6) |
| **LABORATORY:** |  |  |  |
| ASAT | U/L | 10-50 | 66 (22–197) |
| ALAT | U/L | 10-50 | 48 (11–123) |
| Leucocyte | G/L | 4.0 – 10.0 | 9.5 (2.8–25) |
| Lymphocyte | G/L | 1.3-3.6 | 1.1 (0.2–13.2) |
| **RADIOLOGY:** |  |  |  |
| Chest-X-ray | n |  | 30 (44%) |
| Chest-CT-scan |  |  |  |
| - NO CT-scan | n |  | 18 (42%) |
| - Ground Glass | n |  | 19 (46%) |
| - Ground Glass & Consolidation | n |  | 4 (9%) |

Demographic characteristics and blood tests at admission. Continuous measurements were presented as mean (min-max, ±SD) otherwise as median (IQR) if they are normally distributed. Categorical variables were reported as counts and percentages.

### Mechanical ventilation

At admission PaO_2_:FiO_2_ ratios had a median of 87 (54 – 133); with a median FiO_2_ of 72% (60% - 100%) at the ICU admission. Thirty-two (78%) patients received invasive MV, the others were treated with High-Flow Nasal Cannula (Table 2).

Globally, mean PEEP for patients with BMI < 30 Kg/m^2^ was 11 cmH_2_O (10-12, SD 3·80), while mean PEEP for patients with BMI > 30 Kg/m^2^ was 15 cmH_2_O (12-18, SD 3·26). After the onset of MV, the median FiO_2_ improved around 70% (60% – 85%), with first PaO_2_:FiO_2_ ratio with a median of 147 (101 – 233) (Table 2). Thirty-one (75·6%) patients were placed in a prone position (with an average number of pronations of four). In these patients, PaO_2_:FiO_2_ ratio progressively improved during next days, with a median value of 100 (67·5 – 153) during the first day, 142·5 (97·6 – 232·8) during the second day and 167 (113 – 230) during the third day (Table 3).

**Table 2.** Lung starting situation and MV setting.

|  | Unit | n.v. | Results |
| --- | --- | --- | --- |
| **RESPIRATORY DATA:** |  |  |  |
| FiO<sub>2</sub> at admission | % |  | 72 (60–100) |
| P/F-ratio at admission |  | > 300 | 87 (54–133) |
| Respiratory Rate | /min | < 20 | 27 (12–29) |
| FiO<sub>2</sub> after orotracheal intubation | % |  | 70 (60–85) |
| P/F-ratio after orotracheal intubation |  | > 300 | 147 (101–233) |
| **VENTILATORY STRATEGY:** |  |  |  |
| Not-intubated | n |  | 7 (17%) |
| Intubated | n |  | 34 (83%) |
| PEEP-strategy |  |  |  |
| - BMI < 30 Kg/m<sup>2</sup> | cmH<sub>2</sub>O |  | 11 (10–12) - SD 3.8 |
| - BMI > 30 Kg/m<sup>2</sup> | cmH<sub>2</sub>O |  | 15 (12–18) – SD 3.26 |

ICU respiratory data at admission, during treatment and PEEP strategy. Continuous measurements were presented as mean (min-max, ± SD) otherwise as median (IQR) if they are normally distributed. Categorical variables were reported as counts and percentages.

**Table 3.** Clinical evolution.

|  | Unit | n.v. | Results |
| --- | --- | --- | --- |
| **RESPIRATORY DATA:** |  |  |  |
| P/F ratio at first day |  | > 300 | 100.0 (67.5–153.5) |
| P/F ratio at second day |  | > 300 | 142.5 (97.6–232.8) |
| P/F ratio at third day |  | > 300 | 167 (113–230) |
| Pronation cycles | n |  | 4 (1.8–5) |
| **LABORATORY DATA:** |  |  |  |
| C-Reactive-Protein max | mg/L | < 5 | 235.5 (171.0–318.0) |
| Ferritin max | ng/mL | 30-500 | 2225 (1009–3937) |
| Lactate De-Hydrogenase max | U/L | 135-225 | 609 (484–777) |
| Creatinine max | umol/L | 62-106 | 87.5 (75.8–117.5) |
| Troponin T hs max | ng/L |  | 18 (10.9–95.5) |
| Creatinine Kinase max | U/L | 39-308 | 315 (114.3–495.5) |
| Platelets min | G/L | 150-450 | 223.5 (170–305.3) |
| Bilirubin total max | umol/L | < 21.0 | 9.6 (7.8–16.4) |
| **CLINICAL EVOLUTION** |  |  |  |
| Length of hospital stay | days |  | 6.5 (3–10) |
| Length of ICU stay | days |  | 9 (4–12.5) |
| Length of MV | days |  | 6.5 (3–10) |
| **COMPLICATIONS** |  |  |  |
| Ventilator Associated Pneumonia (VAP) | n |  | 8 (19%) |
| Thromboembolism confirmed | n |  | 26 (63%) |
| AKI needing CVVHDF | n |  | 5 (12.2%) |
| **OTHER DRUGS** |  |  |  |
| Tocilizumab | n |  | 20 (49%) |
| Lopinavir/Ritonavir | n |  | 10 (24.4%) |
| Hydroxychloroquine | n |  | 17 (41.4%) |

ICU mechanical ventilation and laboratory data. Continuous measurements were presented as mean (min-max, ± SD) otherwise as median (IQR) if they are normally distributed. Categorical variables were reported as counts and percentages.

### Pharmacologic antithrombotic management

No patient presented any contraindication to be treated with parenteral anticoagulation; 15 (36%) patients were simply treated through prophylaxis, while 26 (63%) patients were managed by full therapeutic anticoagulation (12 – 46% – with Unfractioned Heparin, 14 – 54% – with LMWH). No patient presented any bleeding complication, nor clinical sign requiring anticoagulation reduction or removal.

### ICU patient cohort and survival

The median length of ICU stay was nine days (4 – 12·5); the median duration of MV was seven days (3 – 10) (Table 3). On 12^th^ April, of the 41 patients, ten (24·3%) has died, four (9·7%) are still in the ICU receiving MV (two endotracheal tube, two tracheostomy), two patients were transferred to another hospital and 25 (60.9%) have been discharged from the ICU (Figure 2) in good medical condition with no additional death in the following 10 days. At 7 day from ICU discharge, 39 patients (95%) presented a Karnofsky performance status of more than 80. No patients have been reintubated within or after 72 hours.

**Figure 2.**
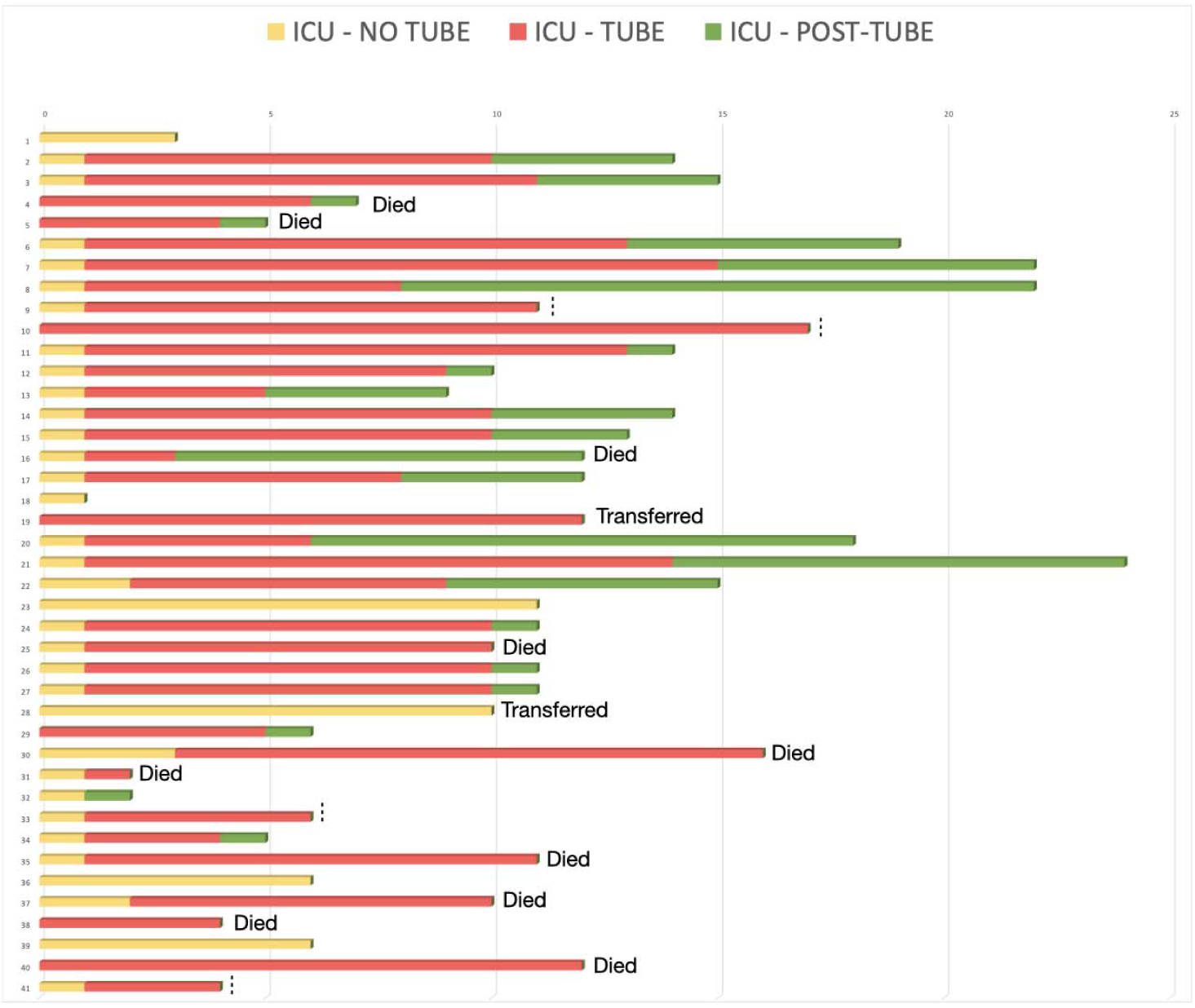
Patients status admitted to the ICU. At April 12^th^, 2020, 10 patients (24.3%) died, 4 (9.7%) are still receiving mechanical ventilation, 2 were transferred to another hospital and 25 (60.9%) were discharged from ICU after a mean of 9 days. All data up to 7 days from extubation have been reported.

### Patients excluded from ICU

The Intensivist Consultant performed clinical counseling of 48 patients admitted to the Internal Medicine Department, ED or from other hospital. The mean age was 65±25 years (38 – 82), presenting SpO_2_ median of 90% (88 – 94), PaO_2_ median of 63·2 mmHg (51·35 – 76·55), a pCO_2_ median of 35·2 mmHg (31·9 – 39·1) and a median value of Hb of 13·9 g/L (12·3 – 15·4). Twenty-six of these patients moved to our ICU and five of them had already been intubated in other hospitals. Intriguingly, the remaining 22 patients consulted but not admitted improved their conditions from an initial median value of SpO_2_ of 87% (84-91%) without being highly symptomatic nor requesting admission in ICU, even for extremely low SpO_2_ values, but with no symptoms of fatigue such as dyspnea (Figure 1).

### Comparison with a published similar cohort

We compared our 41 patients with a cohort of 24 patients published by Bhatraju et al.^18^. Mean age of admission was comparable between the two studies (median 63 ± 12 vs 64 ± 18 yrs), with patients admitted in Lugano having a slightly lower BMI value (29·3 ± 4·94 vs 33·2 ±7·2, p-val 0.007). No significant difference was found in the hematological status of the two cohorts (WBC, Lymphocytes and Platelets) and comparable level of lactate, maximal CRP level during recovery and liver transaminases (Table 1). Median values of the lower P/F ratio were lower in our cohort at day 1 (100 (65 – 162) vs 142 (94-177)), but increased to 142·5 (95·6 – 236·25) and 167 (110·5 – 231) in day two and day three of MV, resulting comparable or higher with the ones reported by Bhatraju et al ^18^.

Overall survival rate during and after ICU admission was longer in our cohort compared to what previously published with a median of 11 days (1-22) versus nine days (1-20) (p-value = 0·028. Figure 3). Length of ICU stay was comparable in both sets (nine days (4 – 12.5) vs nine (4-14) days) but characterized by a lower mortality (10, 24·3%) in our center compared to the 50% mortality reported in Bhatraju et al. ^18^ Median numbers of days of MV was shorter in our cohort (6·5 days (3 – 10)) than what reported in Seattle (8 days (2-20))

**Figure 3.**
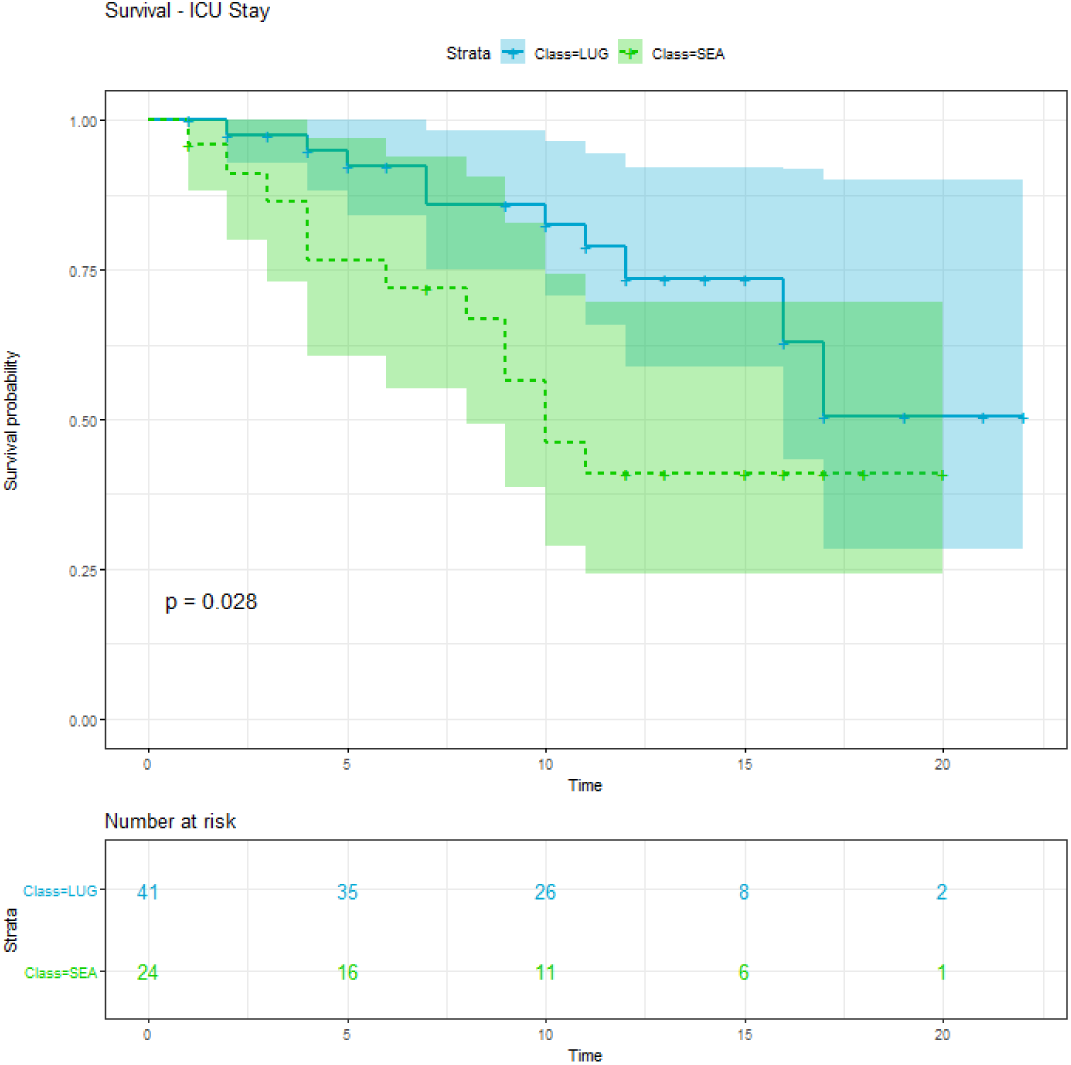
Overall survival rate during and after ICU admission comparison between the study cohort (class LUG; light blue, continuous line) and the data published by Bhatraju et al ^18^ (class SEA; light green, dashed line) (p-value = 0-028).

## DISCUSSION

Acute respiratory distress induced by SARS-CoV-2 is a critical clinical condition associated with the COVID-19 pandemic ^19,20^. It requires adequate preparation in terms of hospital structure, triage systems and clinical training^21^ in order to be correctly addressed and minimize the burden for the patients and the ICU capacity. As COVID-19 is a multisystemic disease, a multidisciplinary approach is mandatory^4^.

To optimize management of tachypnea without dyspnea, also with SpO_2_ lower than normal, surveillance tools such as routine and regular Early Warning Score (EWS) measurement (every 4 hours if stable, reduced hourly if SpO_2_ < 92%)^11,21^ were implemented for intermediate care patients. Patients with COVID-19 interstitial pneumonia present tachypnea correlating with the desaturation degree, without dyspnea or severe neurological symptoms or any organ damage. High pulmonary compliance is probably the responsible for the absence of dyspnea ^22^; we waited until the onset of dyspnea or to a value of 85% of SpO_2_ before admitting any of the patients to the ICU; in such a way, an overload of the ICU was avoided. None of the 48 patients who after a specialist consultation were not admitted to ICU but only placed under clinical surveillance, died in a follow-up from seven up to 21 days. Most of these patients improved their clinical status without being dyspnoeic and some of them have already been discharged from the hospital.

Patients showing a worsening degree of dyspnea transferred to the ICU received “low PEEP ventilatory strategy”, on the contrary to what has been reported in the literature^20^. After intubation we found lungs easy to ventilate, with a higher compliance (on average above 50 mL/cmH_2_O) compared to the “classic ARDS” ^22^. Even if the “classic” criteria for defining the ARDS were confirmed ^23^, there were aspects as the absence of a reduced lung compliance, a “baby-lung” and a consequent tendency to hypercapnia, which induced us to evaluate a more specific treatment, at least in the initial phase.

In according to ARDSnet PEEP table ^13,14^, we preferred to ventilate patients with PEEP tailored to patients’ own BMI, carefully following lungs physiology ^23,24^ This approach would agree with Gattinoni et al ^22^ and Bendjelid et al ^9^, which suggested two different ICU patient populations in COVID-19 pneumonia. The first one presents a high lung compliance and a probable *alveolitis*, with a shunt effect due to loss of local hypoxic vasoconstriction; this population represents the great majority of our patients. The second one presents a low lung compliance and a picture of baby-lung compatible with “classic ARDS” (only two patients in our set).

The “low PEEP ventilatory strategy” we applied allowed us to decrease quickly sedation depth once the inflammation level was reduced. This strategy led to less complications (like ICU paralysis, delayed awakening, agitation, etc..)^18^ and an easier and faster extubation without resorting to large-scale tracheotomy.

During daily screening of the lower limb ultrasound, COVID-19 ICU patients showed a high prevalence rate of DVTs and PEs, even under preventive anticoagulation. In addition, many patients had a marked increase in D-dimers level, partly linked to the finding of DVT and PE in other sites of the body, partly secondary to PE phenomena also on the pulmonary venous side. In this context, it appears reasonable to protect the patient through a pro-active anticoagulant approach than the normal routine. Furthermore, the fact that patients did not encounter any major bleeding phenomenon, supports the idea that in these patients a more aggressive anticoagulation may counterbalance a phenomenon of prothrombotic diathesis, even if the complete mechanism is still unclear.

We observed an increased survival compared to other groups (Figure 2 and 3) ^2,18,24,25^. A possible explanation could be that the relative low-pressure ventilation avoids transforming an initial alveolitis into an ARDS-iatrogenic framework, in which the local ongoing inflammation is rather damaged than helped by high PEEP (generating a Ventilation-Induced-Lung Injury - VILI) in a context that is the “not-classic” ARDS. We observed a very few cases of “classic” ARDS and, in particular, the absence of ARDS cases at the time of admission to the ICU. Mortality in ICU is reported to be as high as 42-62% ^2,24^, while in our dataset is 24.3%. Median days of MV reported by Bhatraju et al. (8 (2-20)) is longer (6·5 (3 – 10)) than what experienced in our clinical setting. In all, this suggests that a less traumatic approach to ventilation by low PEEP and avoiding unnecessary MV by delaying ICU admission can be of help in managing COVID-19 patients and in improving survival.

Our study was burdened by several limitations. First, it was a monocentric observational retrospective study, with a relatively small series of patients. Second, our comparison with current literature is performed on different patient populations, even if cohorts could be considered similar in terms of disease severity and biochemical investigations. This notwithstanding, early data are very encouraging and needs a validation in bigger prospective studies.

In conclusion, the implementation of a multimodal “holistic” approach for COVID-19 patients is highly recommended. We implemented EWS monitoring for intermediate care patients, in order to perform a strict selection of ICU admission and employ MV as little as necessary. MV ventilation was adapted to the real patient needs – i.e. PEEP tailored to patient’s BMI - in order to reduce alveolar traumatism. Anticoagulation screening and therapy has been regulated in order to prevent any sign of thrombosis or thromboembolism. This multimodal program allowed us to reduce the number of ICU admissions, the number of ventilation days and mortality, and could be the base for a further specific patients’ management in this specific contest.

## Data Availability

The data that support the findings of this study are available from the corresponding author, SC, upon reasonable request.

## Declaration of interests

All authors disclose any financial and personal relationships with other people or organizations.

## Author Contributions

S.C., M.R., O.G., M.B. designed the study, performed statistical analysis, and wrote the manuscript; G.B., M.I, PA.M., C.B., S.C., M.C., B.D.M., Z.H., R.L., G.L.P., G.L., D.M., F.O., S.R., S.R., G.T. collected clinical data; C.G., R.M. contributed to study design and interpretation of the data.

## Notes

### Competing Interest Statement

The authors have declared no competing interest.

